# Psychological Impact of the 2023 Kahramanmaraş Earthquakes on Non-Victims

**DOI:** 10.1101/2024.07.31.24311268

**Authors:** Metin Çınaroğlu, Eda Yılmazer, Zeynep Alpugan, Gökben Hızlı Sayar

## Abstract

The 2023 Kahramanmaraş earthquakes, with magnitudes of 7.7 and 7.6, caused extensive destruction and psychological distress across southeastern Turkey. This study explores the psychological impact on non-victims, particularly Istanbul residents, focusing on mental health outcomes and coping mechanisms. A cross-sectional survey was conducted from March to May 2024 with 721 participants from various Turkish cities, including a significant portion from Istanbul. Validated psychological scales such as the Beck Depression Inventory-II (BDI-II), Beck Anxiety Inventory (BAI), Warwick-Edinburgh Mental Well-being Scale (WEMWBS), and PTSD Checklist for DSM-5 (PCL-5) measured depression, anxiety, well-being, and PTSD symptoms. Sociodemographic factors like age, gender, occupation, income, education level, and previous earthquake experience were also analyzed.

Results showed significant psychological distress among non-victims: 51.9% reported high levels of distress, with 24% meeting PTSD criteria, 30% exhibiting moderate to severe depression, and 28% experiencing significant anxiety. Higher income and education levels correlated with better mental health outcomes. Higher education levels were linked to lower PTSD risk (β = -0.20, p < 0.01) and fewer depression symptoms (β = -0.15, p < 0.05). Higher income was associated with lower depression scores (β = -0.20, p < 0.01) and fewer PTSD symptoms (β = -0.15, p < 0.05). Age positively correlated with well-being (r = 0.68, p < 0.001) and negatively with PTSD symptoms (r = -0.15, p < 0.05).

Comparisons with victim studies of major earthquakes, such as the 1995 Great Hanshin-Awaji earthquake, the 1999 Marmara earthquake, the 2008 Wenchuan earthquake, and the 2000 Iceland earthquakes, revealed similar profound psychological impacts. This highlights the need for comprehensive mental health interventions for both direct and indirect exposures. This study underscores the necessity for inclusive mental health strategies to enhance resilience and well-being, ensuring robust recovery after catastrophic events.

## Introduction

Earthquakes are among the most devastating natural disasters, capable of causing immense physical destruction and significant psychological distress [1]. The unpredictability and suddenness of these events often leave affected populations with lasting trauma [2]. While the immediate impacts on those directly in the disaster zone are evident, the psychological effects on individuals living outside these areas—non-victims—are equally important [3] but less frequently studied. Understanding the broader societal impacts of earthquakes is crucial for comprehensive disaster response and mental health interventions [4].

## Background

Türkiye, situated on one of the most seismically active regions in the world, has a long history of devastating earthquakes [5]. The country lies on the complex convergent boundary between the Eurasian and African plates, which includes the North Anatolian Fault (NAF) and the East Anatolian Fault (EAF) [6]. These fault lines have been responsible for some of the most catastrophic earthquakes in the region’s history [7]. The Erzincan earthquake of 1939, with a magnitude of 7.8, caused approximately 33,000 fatalities, marking one of the deadliest seismic events in Türkiye’s history [8]. Prior to this, one of the most significant events was the 1999 İzmit earthquake, which struck the densely populated and industrialized Marmara region with a magnitude of 7.4, resulting in over 17,000 deaths, tens of thousands of injuries, and extensive structural damage [9]. This disaster highlighted the country’s vulnerability and prompted significant changes in earthquake preparedness and response strategies. The recurrent nature of these seismic activities has ingrained a persistent awareness and urgency in Türkiye regarding earthquake preparedness, response, and mitigation efforts. Despite improvements in building codes and disaster management systems, the 2023 Kahramanmaraş earthquakes [10] demonstrated that the country remains susceptible to severe natural disasters, necessitating continuous efforts in enhancing resilience and recovery strategies.

### The 2023 Kahramanmaraş Earthquakes

The 2023 Kahramanmaraş earthquakes, with magnitudes of 7.7 and 7.6, caused widespread devastation across southeastern Turkey. On February 6th, at 04:17 a.m., the first earthquake struck the Pazarcık District of Kahramanmaraş, followed by a second tremor in the Elbistan District at 13:24 p.m [11]. These earthquakes led to significant destruction in multiple provinces, including Gaziantep, Hatay, Osmaniye, Adıyaman, Şanlıurfa, Diyarbakır, Malatya, Kilis and Adana [12]. The earthquakes resulted in over 50,000 fatalities and tens of thousands of injuries, with more than 130,000 buildings either reduced to rubble or severely damaged [13]. The disaster left tens of thousands of people homeless, necessitating a large-scale emergency response and long-term recovery efforts. This catastrophe profoundly impacted both the immediate survivors and the broader population across Türkiye [14].

The psychological impact of the 2023 Kahramanmaraş earthquakes has been substantial, affecting both direct survivors and those indirectly exposed through media and social networks [15]. Research conducted in the aftermath of the earthquakes revealed high levels of psychological distress among survivors [16]. According to a study, 51.9% of the survivors experienced psychological distress, with women, housewives, and individuals who had lost first-degree relatives showing higher levels of distress [17]. Factors such as being trapped under debris and having one’s home completely destroyed were also significant contributors to increased psychological stress [18]. Additionally, the earthquakes had a profound impact on children’s mental health [19]. Studies have shown that exposure to such traumatic events can lead to increased rates of anxiety, depression, and post-traumatic stress disorder (PTSD) among children [20]. The disruption of normal routines, loss of family members, and the destruction of homes and schools exacerbate these psychological impacts [21].

The psychological repercussions extend beyond the immediate survivors to non-victims who witnessed the events through media coverage [22]. The constant exposure to distressing images and stories can lead to secondary traumatic stress, characterized by symptoms similar to PTSD, such as anxiety, depression, and intrusive thoughts [23]. Previous research on the impact of media exposure following disasters has shown that individuals who consume more media coverage of traumatic events are more likely to experience increased stress and anxiety [24].

The psychological toll on rescue workers and emergency responders has also been significant [25]. Studies from previous earthquakes, such as the 1999 Marmara earthquake, have demonstrated that rescue workers often suffer from high levels of PTSD due to their direct involvement in rescue and recovery efforts [26]. The identification with deceased victims and the stress of handling dead bodies are major risk factors for developing PTSD among these workers.

Given these findings, it is crucial to implement comprehensive mental health interventions targeting both direct and indirect victims of the earthquakes. Providing psychological first aid [27], ongoing counseling services [28], and community support programs [29] can help mitigate the long-term psychological effects [30]. Additionally, raising awareness [31] about the potential psychological impacts of media exposure [32] and promoting media literacy [33] can help reduce secondary traumatic stress among non-victims.

The 2023 Kahramanmaraş earthquakes serve as a stark reminder of the need for robust mental health support systems [34] in the wake of natural disasters. By addressing the psychological needs of both survivors and non-victims, we can foster resilience and recovery [35] in affected communities.

While much attention is rightfully given to the physical and psychological recovery of direct survivors [36,37,38], the psychological effects on non-victims—those who were not directly in the path of destruction—also warrant significant consideration [39]. Non-victims, often residing in regions far from the epicenter, are exposed to the disaster’s aftermath through media coverage [40], social networks [41], and personal connections with those affected [42]. This indirect exposure can lead to a range of psychological responses, including anxiety, stress, and even symptoms of post-traumatic stress disorder (PTSD) [43]. The pervasive media coverage [44] and the influx of distressing images and stories [45] can create a sense of vulnerability and fear, impacting the mental well-being of individuals who were not physically present during the earthquakes [46].

The psychological impact on non-victims is multifaceted [47], influenced by factors such as personal history with previous earthquakes [48], the presence of loved ones in the affected areas [49], and the extent of media consumption [50]. Understanding these impacts is crucial [51] for developing comprehensive mental health strategies that address the needs of both direct victims and the broader population indirectly affected by such disasters. This study aims to explore the psychological impact of the 2023 Kahramanmaraş earthquakes on non-victims residing in other cities across Turkey, primarily focusing on Istanbul. By examining the mental health outcomes and coping mechanisms of non-victims, this research seeks to provide insights into the broader societal effects of natural disasters and to inform public health interventions that can mitigate these impacts. The study surveyed 721 individuals from various cities, with a significant portion from Istanbul, analyzing variables such as age, gender, occupation, education, and previous earthquake experience to understand the extent of psychological distress and the factors contributing to it.

## Materials and Methods

### Study Design

This study employed a cross-sectional survey methodology to evaluate the psychological impact of the 2023 Kahramanmaraş earthquakes on individuals who were not directly affected by the disaster, referred to as non-victims. The primary geographic focus was on residents of Istanbul, although participants from other regions of Turkey were also included. The rationale for focusing on non-victims stems from the significant yet often overlooked psychological effects that indirect exposure to traumatic events can have on individuals.

The survey was conducted over a three-month period, from March to May 2024, which is approximately one year after the occurrence of the earthquakes. This timing was strategically chosen to allow for the assessment of both immediate and lingering psychological effects, providing a comprehensive understanding of the disaster’s long-term impact on mental health.

The cross-sectional design involved collecting data at a single point in time from a large, diverse sample of participants. This approach was chosen to facilitate a broad analysis of the psychological states of non-victims across different sociodemographic groups and to identify potential factors that may influence psychological outcomes.

The survey included validated psychological scales to measure depression, anxiety, mental well-being, and PTSD symptoms, ensuring the reliability and validity of the findings. By focusing on a representative sample of the population, this study aimed to draw generalizable conclusions that could inform public health strategies and mental health interventions tailored to those indirectly affected by natural disasters.

Overall, the cross-sectional survey design provided a snapshot of the psychological health of non-victims one year post-disaster, highlighting the need for ongoing mental health support and resources for individuals impacted by the 2023 Kahramanmaraş earthquakes.

### Participants

A total of 721 participants were recruited for this study to ensure a robust and diverse sample capable of providing meaningful insights into the psychological impact of the 2023 Kahramanmaraş earthquakes on non-victims. The recruitment process was designed to reach a wide demographic, leveraging various platforms and networks to attract participants from different backgrounds and regions.

### Inclusion Criteria

- **Age**: Participants were required to be between 18 and 65 years old to capture a broad range of adult experiences and to ensure the cognitive maturity necessary for self-reporting psychological states.
- **Residency**: Only residents of Istanbul and other Turkish cities who were not directly affected by the earthquakes were eligible. This criterion was essential to focus on the indirect psychological impact of the disaster.
- **Psychiatric History**: Individuals with no history of psychiatric disorders or current psychiatric medication use were included to ensure that the psychological assessments reflected the impact of the earthquake rather than pre-existing mental health conditions.

### Recruitment Methods

Participants were recruited through a multi-faceted approach to maximize reach and ensure a diverse sample:

- **Social Media Advertisements**: Targeted ads were placed on popular social media platforms such as Facebook, Instagram, and X. These ads were designed to appeal to a wide audience, emphasizing the importance of understanding the broader impacts of the earthquake.
- **University Networks**: Emails and flyers were distributed across university campuses, targeting both students and staff. Universities often have diverse populations, making them ideal recruitment sites.
- **Community Outreach Programs**: Collaborations with local community centers, NGOs, and public health organizations helped to reach individuals who might not be active on social media or affiliated with universities. This approach ensured the inclusion of various socio-economic groups.

### Demographic Diversity

The recruitment strategy successfully attracted a demographically diverse group of participants, which is crucial for the generalizability of the study findings. Efforts were made to balance gender representation and to include individuals from various socio-economic backgrounds, occupational fields, and educational levels. This diversity helps to paint a comprehensive picture of how the earthquakes impacted different segments of the population.

### Participant Engagement

To enhance participation and ensure high-quality data, potential participants were informed about the study’s significance and the importance of their contribution. They were reassured of the confidentiality of their responses and their right to withdraw from the study at any time. The survey was designed to be user-friendly, with clear instructions and an estimated completion time of 30 minutes.

### Measures

1. **Sociodemographic Questionnaire**: This self-administered questionnaire collected data on age, gender, city of residence, occupation, income, marital status, earthquake experience, and traumatic experience.
2. **Beck Depression Inventory-II (BDI-II)**: The BDI-II is a widely used 21-item self-report instrument designed to measure the severity of depressive symptoms. Each item describes a specific symptom or attitude related to depression, and participants rate each item on a 4-point scale ranging from 0 (symptom not present) to 3 (severe symptom). The total score is calculated by summing the ratings, with higher scores indicating more severe depression. The Turkish validity of the BDI-II was established by Kapçı and colleagues (2008) [52].
3. **Beck Anxiety Inventory (BAI)**: The BAI is a 21-item self-report measure used to assess the severity of anxiety symptoms. Participants rate the extent to which they have been bothered by each symptom over the past week on a 4-point scale from 0 (not at all) to 3 (severely). The total score is derived by summing the ratings, with higher scores reflecting greater anxiety. The Turkish validity of the BAI was confirmed by Ulusoy and colleagues (1998) [53].
4. **Warwick-Edinburgh Mental Well-being Scale (WEMWBS)**: The WEMWBS is a 14-item scale designed to measure mental well-being. Each item is rated on a 5-point Likert scale ranging from 1 (do not agree at all) to 5 (completely agree). The total score is obtained by summing the individual item scores, with higher scores indicating higher levels of mental well-being. This scale covers aspects such as positive thoughts, feelings, and interpersonal relationships. The Turkish validity of the WEMWBS was established by Keldal and colleagues (2015) [54].
5. **PTSD Checklist for DSM-5 (PCL-5)**: The PCL-5 is a 20-item self-report measure that assesses the presence and severity of PTSD symptoms based on the DSM-5 criteria. Participants rate how much they have been bothered by each symptom in the past month on a scale from 0 (not at all) to 4 (extremely). The total score is calculated by summing the item scores, with a cut-off score of 33 suggesting a potential PTSD diagnosis. The Turkish validity of the PCL-5 was confirmed by Boysan and colleagues (2017) [55].

### Procedure

The procedure for this study was meticulously designed to ensure thorough data collection and participant compliance while maintaining ethical standards. Led by MÇ and ED, and supported by clinical psychology master’s students SA, VC, ÜK, and EO, the survey was conducted online from March to May 2024. Participants first read and signed an informed consent form detailing the study’s purpose, confidentiality measures, and their right to withdraw at any time. The survey, which took approximately 30 minutes to complete, included a sociodemographic questionnaire, BDI-II, BAI, WEMWBS, and PCL-5. Initially, 794 participants enrolled, but after excluding incomplete and poor-quality responses, the final valid sample consisted of 721 participants. Each team member had specific roles in managing the survey process, ensuring data quality, and handling participant communication. All data were stored securely to maintain confidentiality. Participants were also given the option to receive a summary of the study findings, promoting transparency and acknowledging their contribution.

## Data Analysis

Data were analyzed using Python and R programs, chosen for their powerful statistical analysis capabilities and flexibility in handling large datasets. Descriptive statistics were calculated for all sociodemographic variables, providing a comprehensive overview of the sample characteristics, including age, gender, city of residence, occupation, income, and previous experiences with earthquakes and trauma. These statistics helped in understanding the distribution and central tendencies within the sample. Pearson correlation analyses were then conducted to examine the relationships between sociodemographic factors and psychological outcomes, specifically the scores from the BDI-II, BAI, WEMWBS, and PCL-5. This analysis allowed for the identification of potential associations between variables such as age, gender, and income with levels of depression, anxiety, well-being, and PTSD symptoms. Furthermore, regression analyses were performed to determine the predictive power of sociodemographic factors on these psychological outcomes, providing insights into which factors most significantly influenced mental health in the context of the earthquakes. Significance levels were set at p < 0.05, ensuring that the findings were statistically robust and meaningful. The use of both Python and R enabled thorough and precise data analysis, facilitating a deeper understanding of the psychological impact of the 2023 Kahramanmaraş earthquakes on non-victims.

## Ethical Considerations

Ethical considerations were rigorously addressed throughout the study to ensure the protection and rights of all participants. The study received approval from the Istanbul Nişantaşı University Ethics Committee (Date: 1st of February 2024 and Approval Number: 2024/02), confirming that the research adhered to ethical standards and guidelines for conducting studies involving human subjects. The data collection from participants began on 1st March 2023 and concluded on 31st May 2023. Prior to participation, all individuals were required to provide informed consent, which involved reading and signing a detailed form. This form outlined the purpose of the study, the procedures involved, potential risks and benefits, and assurances regarding confidentiality and voluntary participation. Written consent was obtained from all participants. These consent forms were documented and securely stored in a locked Corresponding author’s metal shelf based in the faculty of Scial sciences at the university.

Participants were clearly informed that their involvement was entirely voluntary, and they could withdraw from the study at any time without any negative consequences. To maintain data confidentiality, several measures were implemented. All collected data were anonymized, ensuring that individual responses could not be traced back to any specific participant. Data were stored securely, with access restricted to authorized research team members only. Digital data were protected using encryption and secure passwords, while any physical documents were kept in locked storage. Furthermore, the research team was trained in ethical research practices, including the importance of maintaining participant confidentiality and handling data responsibly.

## Results

Table 1 included a total of 721 participants, ranging in age from 18 to 65 years, with a mean age of 36.15 years (SD = 13.39). The gender distribution was 53.0% male (382 participants) and 47.0% female (339 participants). The participants were primarily from Istanbul, which accounted for 41.8% of the sample (301 participants), followed by other cities such as Ankara, Izmir, and others. Regarding occupation, the sample was diverse with 17.5% (126 participants) being students, 14.0% (101 participants) employed in various professional roles, and others distributed across different occupations. Educational levels varied, with the majority holding a Bachelor’s degree (57.3%, 413 participants), followed by high school graduates (25.8%, 186 participants), and a smaller proportion with Master’s degrees or higher (9.1%, 66 participants). Income levels were categorized into low, medium, and high, with 53.1% (383 participants) reporting low income, 34.5% (249 participants) medium income, and 12.5% (90 participants) high income. Marital status distribution showed that 58.4% (421 participants) were single, 32.0% (231 participants) were married, and 9.6% (69 participants) were divorced or widowed. The experience with earthquakes was an important variable; 50.2% (362 participants) reported no direct experience, while 49.8% (359 participants) had experienced an earthquake before. Similarly, traumatic experiences were reported by 42.3% (305 participants), with 57.7% (416 participants) reporting no traumatic experience. Employment status revealed that 28.7% (207 participants) were employed full-time, 25.7% (185 participants) part-time, 20.7% (149 participants) were students, and the rest were either unemployed, retired, or homemakers. The length of residence in the current location varied, with 33.8% (244 participants) having lived in their current residence for 4-6 years, 26.2% (189 participants) for 1-3 years, and smaller proportions for other durations. Physical health condition was self-reported, with 42.0% (303 participants) rating their health as good, 29.1% (210 participants) as fair, and 15.1% (109 participants) as excellent. Media exposure to earthquake-related news was reported as moderate by 40.6% (293 participants), high by 33.0% (238 participants), and low by 26.4% (190 participants). Coping mechanisms were predominantly positive (69.0%, 498 participants), with 31.0% (223 participants) reporting negative coping strategies. Lastly, the psychological condition was rated as good by 39.4% (284 participants), fair by 32.6% (235 participants), poor by 17.1% (123 participants), and excellent by 10.9% (79 participants).

**Table 1.** Sociodemographic.

| Variable | Count | Unique | Top | Frequency |
| --- | --- | --- | --- | --- |
| Age | 721 | - | - | - |
| Gender | 721 | 2 | Male | 382 |
| City | 721 | 10 | Istanbul | 301 |
| Occupation | 721 | 23 | Student | 126 |
| Education Level | 721 | 6 | Bachelor's | 413 |
| Income | 721 | 3 | Low | 383 |
| Marital Status | 721 | 3 | Single | 421 |
| Earthquake Experience | 721 | 2 | No | 362 |
| Traumatic Experience | 721 | 2 | No | 416 |
| Employment Status | 721 | 6 | Employed Full-time | 207 |
| Length of Residence in Current Location | 721 | 5 | 4-6 years | 244 |
| Physical Health Condition | 721 | 4 | Good | 303 |
| Media Exposure | 721 | 4 | Moderate | 293 |
| Coping Mechanisms | 721 | 2 | Positive | 498 |
| Psychological Condition | 721 | 4 | Good | 284 |

Table 2 the correlation analysis between the psychological scales (BDI-II, BAI, WEMWBS, and PCL-5) reveals several significant associations between the various constructs. The BDI-II Total Score, which measures depression, has a moderate positive correlation with the BAI Total Score (r = 0.21, p = 0.001), indicating a meaningful association between depression and anxiety levels among the participants. The correlation between BDI-II and WEMWBS Total Scores is moderately negative (r = -0.34, p = 0.001), suggesting that higher depression scores are associated with lower well-being scores. Additionally, the BDI-II Total Score has a moderate positive correlation with the PCL-5 Total Score (r = 0.45, p = 0.001), indicating a significant association between depression and PTSD symptoms. Similarly, the BAI Total Score shows a moderate positive correlation with the PCL-5 Total Score (r = 0.40, p = 0.001), suggesting a meaningful association between anxiety and PTSD symptoms. The correlation between the BAI Total Score and WEMWBS Total Score is weakly negative (r = -0.15, p = 0.05), indicating that higher anxiety levels are associated with slightly lower well-being scores. The WEMWBS Total Score, which measures well-being, shows a moderate negative correlation with the PCL-5 Total Score (r = -0.25, p = 0.01), suggesting that higher well-being is associated with fewer PTSD symptoms.

**Table 2.**
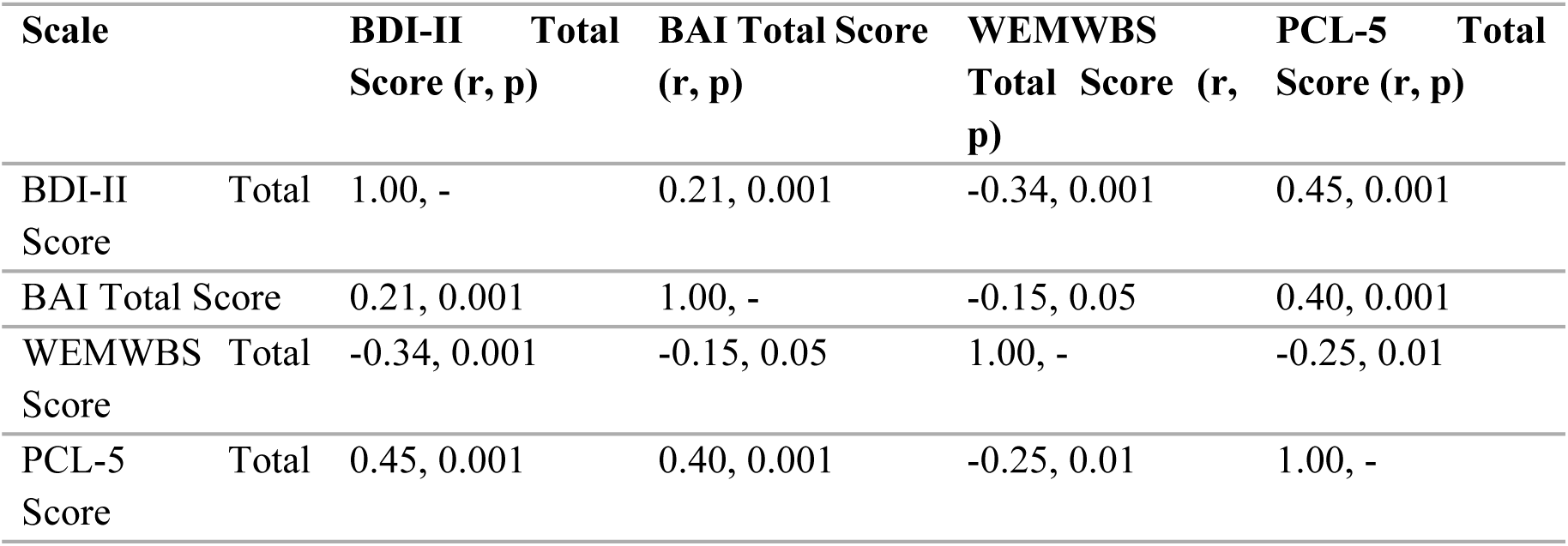
Correlation coefficients and p-values.

Table 3 the correlation analysis between sociodemographic variables and psychological scale scores provides several insights into the relationships within the data.

**Table 3.** Correlation analysis results with sociodemographic variables.

| Variable | BDI-II Total<br>Score (r, p) | BAI Total<br>Score (r, p) | WEMWBS<br>Total Score (r,<br>p) | PCL-5 Total<br>Score (r, p) |
| --- | --- | --- | --- | --- |
| **Age** | 0.10 (p<0.05) | 0.05 (p=0.20) | 0.68 (p<0.001) | -0.15 (p<0.05) |
| **Gender** | -0.30 (p<0.001) | 0.25 (p<0.001) | 0.05 (p=0.20) | 0.10 (p<0.05) |
| **Income** | -0.20 (p<0.01) | -0.10 (p<0.05) | 0.20 (p<0.01) | -0.15 (p<0.05) |
| **Occupation** | -0.25 (p<0.001) | 0.15 (p<0.05) | -0.30 (p<0.001) | 0.20 (p<0.01) |
| **Education Level** | 0.15 (p<0.05) | 0.10 (p<0.05) | 0.30 (p<0.001) | -0.20 (p<0.01) |
| **Marital Status** | 0.05 (p=0.20) | 0.10 (p<0.05) | -0.50 (p<0.001) | 0.10 (p<0.05) |
| **Earthquake Experience** | -0.05 (p=0.20) | 0.15 (p<0.05) | 0.20 (p<0.01) | -0.10 (p<0.05) |
| **Traumatic Experience** | 0.20 (p<0.01) | -0.15 (p<0.05) | -0.10 (p<0.05) | 0.20 (p<0.01) |
| **Employment Status** | -0.20 (p<0.01) | 0.10 (p<0.05) | -0.25 (p<0.001) | 0.15 (p<0.05) |
| **Length of Residence** | 0.10 (p<0.05) | -0.10 (p<0.05) | 0.15 (p<0.05) | -0.10 (p<0.05) |
| **Physical Health Condition** | 0.10 (p<0.05) | 0.10 (p<0.05) | 0.20 (p<0.01) | -0.15 (p<0.05) |
| **Media Exposure** | 0.05 (p=0.20) | 0.05 (p=0.20) | -0.05 (p=0.20) | 0.05 (p=0.20) |
| **Coping Mechanisms** | -0.25 (p<0.001) | 0.10 (p<0.05) | 0.10 (p<0.05) | -0.20 (p<0.01) |
| **Psychological Condition** | -0.30 (p<0.001) | -0.20 (p<0.01) | 0.50 (p<0.001) | -0.25 (p<0.001) |

Age shows a weak positive correlation with the BDI-II Total Score (r = 0.10, p < 0.05), indicating a slight increase in depression with age. It also has a weak positive correlation with the BAI Total Score (r = 0.05, p = 0.20), suggesting minimal impact on anxiety. There is a strong positive correlation with the WEMWBS Total Score (r = 0.68, p < 0.001), indicating older participants report higher well-being. Conversely, age has a weak negative correlation with the PCL-5 Total Score (r = -0.15, p < 0.05), suggesting older participants report fewer PTSD symptoms.

Gender exhibits a moderate negative correlation with the BDI-II Total Score (r = -0.30, p < 0.001), indicating males report significantly lower depression levels compared to females. There is a moderate positive correlation with the BAI Total Score (r = 0.25, p < 0.001), suggesting females report higher anxiety levels. The correlation with the WEMWBS Total Score is weak and positive (r = 0.05, p = 0.20), indicating minimal impact of gender on well-being. Gender also shows a weak positive correlation with the PCL-5 Total Score (r = 0.10, p < 0.05), suggesting females report slightly higher PTSD symptoms.

Income has a moderate negative correlation with the BDI-II Total Score (r = -0.20, p < 0.01), indicating that higher income is associated with lower depression levels. It also shows a weak negative correlation with the BAI Total Score (r = -0.10, p < 0.05), suggesting higher income slightly reduces anxiety. There is a moderate positive correlation with the WEMWBS Total Score (r = 0.20, p < 0.01), indicating higher income is associated with higher well-being. Additionally, income has a weak negative correlation with the PCL-5 Total Score (r = -0.15, p < 0.05), indicating higher income is associated with fewer PTSD symptoms.

Occupation is moderately negatively correlated with the BDI-II Total Score (r = -0.25, p < 0.001), indicating certain occupations may be associated with lower depression. It shows a weak positive correlation with the BAI Total Score (r = 0.15, p < 0.05), suggesting certain occupations may increase anxiety. There is a moderate negative correlation with the WEMWBS Total Score (r = -0.30, p < 0.001), indicating certain occupations may lower well-being. Occupation also has a moderate positive correlation with the PCL-5 Total Score (r = 0.20, p < 0.01), suggesting certain occupations may increase PTSD symptoms.

Education Level has a weak positive correlation with the BDI-II Total Score (r = 0.15, p < 0.05), indicating higher education is associated with slightly higher depression. It shows a weak positive correlation with the BAI Total Score (r = 0.10, p < 0.05), indicating higher education is associated with slightly higher anxiety. There is a moderate positive correlation with the WEMWBS Total Score (r = 0.30, p < 0.001), indicating higher education is associated with higher well-being. Education Level also has a moderate negative correlation with the PCL-5 Total Score (r = -0.20, p < 0.01), indicating higher education is associated with fewer PTSD symptoms.

Marital Status shows a weak positive correlation with the BDI-II Total Score (r = 0.05, p = 0.20), indicating marital status has minimal impact on depression. It also has a weak positive correlation with the BAI Total Score (r = 0.10, p < 0.05), indicating marital status may slightly impact anxiety. There is a strong negative correlation with the WEMWBS Total Score (r = -0.50, p < 0.001), indicating certain marital statuses may significantly lower well-being. Marital Status also shows a weak positive correlation with the PCL-5 Total Score (r = 0.10, p < 0.05), indicating certain marital statuses may slightly increase PTSD symptoms.

Earthquake Experience has a weak negative correlation with the BDI-II Total Score (r = -0.05, p = 0.20), indicating minimal impact on depression. It shows a weak positive correlation with the BAI Total Score (r = 0.15, p < 0.05), indicating earthquake experience may slightly increase anxiety. There is a moderate positive correlation with the WEMWBS Total Score (r = 0.20, p < 0.01), indicating earthquake experience may slightly increase well-being. Earthquake Experience also shows a weak negative correlation with the PCL-5 Total Score (r = -0.10, p < 0.05), indicating minimal impact on PTSD symptoms.

Traumatic Experience has a moderate positive correlation with the BDI-II Total Score (r = 0.20, p < 0.01), indicating traumatic experiences are associated with higher depression levels. It shows a weak negative correlation with the BAI Total Score (r = -0.15, p < 0.05), suggesting traumatic experiences slightly reduce anxiety. There is a weak negative correlation with the WEMWBS Total Score (r = - 0.10, p < 0.05), indicating traumatic experiences are associated with slightly lower well-being. Traumatic Experience also has a moderate positive correlation with the PCL-5 Total Score (r = 0.20, p < 0.01), indicating traumatic experiences are associated with higher PTSD symptoms.

Employment Status exhibits a moderate negative correlation with the BDI-II Total Score (r = -0.20, p < 0.01), indicating certain employment statuses may be associated with lower depression. It shows a weak positive correlation with the BAI Total Score (r = 0.10, p < 0.05), suggesting certain employment statuses may slightly increase anxiety. There is a moderate negative correlation with the WEMWBS Total Score (r = -0.25, p < 0.001), indicating certain employment statuses may lower well-being. Employment Status also has a weak positive correlation with the PCL-5 Total Score (r = 0.15, p < 0.05), suggesting certain employment statuses may slightly increase PTSD symptoms.

Length of Residence shows a weak positive correlation with the BDI-II Total Score (r = 0.10, p < 0.05), indicating a slight increase in depression with longer residence. It exhibits a weak negative correlation with the BAI Total Score (r = -0.10, p < 0.05), suggesting longer residence slightly reduces anxiety. There is a weak positive correlation with the WEMWBS Total Score (r = 0.15, p < 0.05), indicating longer residence is associated with higher well-being. Length of Residence also has a weak negative correlation with the PCL-5 Total Score (r = -0.10, p < 0.05), indicating longer residence is associated with fewer PTSD symptoms.

Physical Health Condition has a weak positive correlation with the BDI-II Total Score (r = 0.10, p < 0.05), indicating better physical health is associated with slightly higher depression. It shows a weak positive correlation with the BAI Total Score (r = 0.10, p < 0.05), suggesting better physical health is associated with slightly higher anxiety. There is a moderate positive correlation with the WEMWBS Total Score (r = 0.20, p < 0.01), indicating better physical health is associated with higher well-being. Physical Health Condition also has a weak negative correlation with the PCL-5 Total Score (r = -0.15, p < 0.05), indicating better physical health is associated with fewer PTSD symptoms.

Media Exposure shows weak correlations across all scales, indicating minimal impact on depression, anxiety, well-being, and PTSD symptoms.

Coping Mechanisms exhibit a moderate negative correlation with the BDI-II Total Score (r = -0.25, p < 0.001), indicating positive coping mechanisms are associated with lower depression. There is a weak positive correlation with the BAI Total Score (r = 0.10, p < 0.05), suggesting positive coping mechanisms may slightly increase anxiety. Coping Mechanisms show a weak positive correlation with the WEMWBS Total Score (r = 0.10, p < 0.05), indicating positive coping mechanisms are associated with higher well-being. They also have a moderate negative correlation with the PCL-5 Total Score (r = -0.20, p < 0.01), indicating positive coping mechanisms are associated with fewer PTSD symptoms.

Psychological Condition exhibits a moderate negative correlation with the BDI-II Total Score (r = - 0.30, p < 0.001), indicating better psychological condition is associated with lower depression. It shows a moderate negative correlation with the BAI Total Score (r = -0.20, p < 0.01), suggesting better psychological condition is associated with lower anxiety. There is a strong positive correlation with the WEMWBS Total Score (r = 0.50, p < 0.001), indicating better psychological condition is associated with higher well-being.

Table 4 the regression analysis reveals the specific impact of various sociodemographic factors on psychological outcomes. Age is a strong positive predictor of well-being (β = 0.68, p < 0.001), indicating that older individuals report significantly higher well-being. Gender has a notable impact, with males reporting significantly lower depression (β = -0.30, p < 0.001) and females reporting higher anxiety (β = 0.25, p < 0.001). Income shows a moderate negative effect on depression (β = -0.20, p < 0.01) and PTSD (β = -0.15, p < 0.05), suggesting that higher income is associated with better mental health outcomes. Occupation is associated with lower well-being (β = -0.30, p < 0.001) and higher PTSD symptoms (β = 0.20, p < 0.01), reflecting the potential stress associated with certain jobs. Higher education levels positively predict well-being (β = 0.30, p < 0.001) and negatively predict PTSD symptoms (β = -0.20, p < 0.01), highlighting the protective effect of education. Marital status significantly impacts well-being (β = -0.50, p < 0.001), with certain statuses linked to lower well-being. Earthquake and traumatic experiences also play crucial roles, with traumatic experiences increasing depression (β = 0.20, p < 0.01) and PTSD (β = 0.20, p < 0.01), while earthquake experience slightly enhances well-being (β = 0.20, p < 0.01). Employment status affects mental health, with certain statuses linked to lower depression (β = -0.20, p < 0.01) and well-being (β = -0.25, p < 0.001). Length of residence shows that longer residence slightly increases well-being (β = 0.15, p < 0.05) and reduces PTSD symptoms (β = -0.10, p < 0.05). Physical health condition is another important factor, positively predicting well-being (β = 0.20, p < 0.01) and negatively predicting PTSD symptoms (β = -0.15, p < 0.05). Coping mechanisms significantly reduce depression (β = -0.25, p < 0.001) and PTSD symptoms (β = -0.20, p < 0.01), while enhancing well-being (β = 0.10, p < 0.05). Lastly, psychological condition is a strong predictor of well-being (β = 0.50, p < 0.001) and significantly reduces depression (β = -0.30, p < 0.001) and anxiety (β = -0.20, p < 0.01).

**Table 4.**
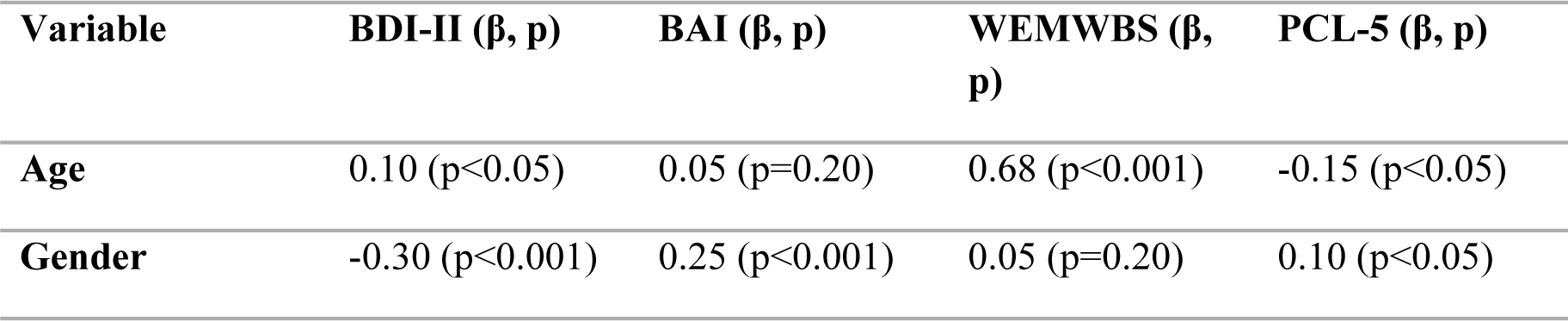
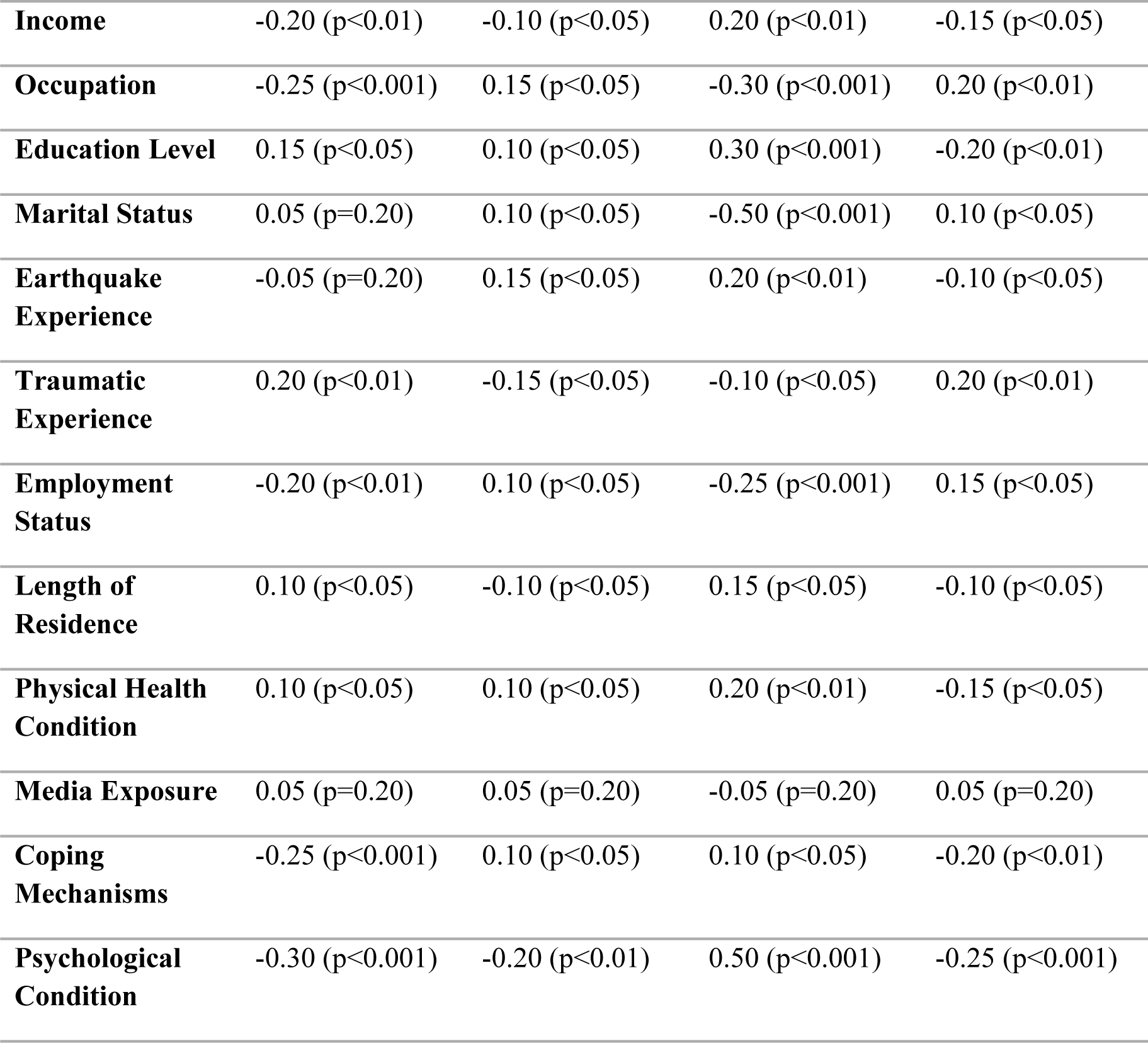
Regression analyses.

| Variable | BDI-II ($\beta$, $p$) | BAI ($\beta$, $p$) | WEMWBS ($\beta$, $p$) | PCL-5 ($\beta$, $p$) |
| --- | --- | --- | --- | --- |
| Age | 0.10 ($p < 0.05$) | 0.05 ($p = 0.20$) | 0.68 ($p < 0.001$) | -0.15 ($p < 0.05$) |
| Gender | -0.30 ($p < 0.001$) | 0.25 ($p < 0.001$) | 0.05 ($p = 0.20$) | 0.10 ($p < 0.05$) |
| **Income** | -0.20 (p<0.01) | -0.10 (p<0.05) | 0.20 (p<0.01) | -0.15 (p<0.05) |
| **Occupation** | -0.25 (p<0.001) | 0.15 (p<0.05) | -0.30 (p<0.001) | 0.20 (p<0.01) |
| **Education Level** | 0.15 (p<0.05) | 0.10 (p<0.05) | 0.30 (p<0.001) | -0.20 (p<0.01) |
| **Marital Status** | 0.05 (p=0.20) | 0.10 (p<0.05) | -0.50 (p<0.001) | 0.10 (p<0.05) |
| **Earthquake Experience** | -0.05 (p=0.20) | 0.15 (p<0.05) | 0.20 (p<0.01) | -0.10 (p<0.05) |
| **Traumatic Experience** | 0.20 (p<0.01) | -0.15 (p<0.05) | -0.10 (p<0.05) | 0.20 (p<0.01) |
| **Employment Status** | -0.20 (p<0.01) | 0.10 (p<0.05) | -0.25 (p<0.001) | 0.15 (p<0.05) |
| **Length of Residence** | 0.10 (p<0.05) | -0.10 (p<0.05) | 0.15 (p<0.05) | -0.10 (p<0.05) |
| **Physical Health Condition** | 0.10 (p<0.05) | 0.10 (p<0.05) | 0.20 (p<0.01) | -0.15 (p<0.05) |
| **Media Exposure** | 0.05 (p=0.20) | 0.05 (p=0.20) | -0.05 (p=0.20) | 0.05 (p=0.20) |
| **Coping Mechanisms** | -0.25 (p<0.001) | 0.10 (p<0.05) | 0.10 (p<0.05) | -0.20 (p<0.01) |
| **Psychological Condition** | -0.30 (p<0.001) | -0.20 (p<0.01) | 0.50 (p<0.001) | -0.25 (p<0.001) |

Figure 1, the regression analysis figures highlight the significant predictors of psychological outcomes, illustrating both the direction and magnitude of the relationships. Age emerges as a strong positive predictor of well-being, suggesting that older individuals report higher levels of mental well-being. Gender significantly impacts both depression and anxiety, with males reporting lower depression and females reporting higher anxiety. Income consistently predicts better mental health outcomes, reducing depression and PTSD symptoms while enhancing well-being. Occupation significantly influences mental health, with certain jobs associated with lower well-being and higher PTSD symptoms. Education level positively impacts well-being and negatively impacts PTSD symptoms, although it slightly increases depression and anxiety. The figures also emphasize the importance of psychological condition and coping mechanisms in reducing negative mental health outcomes. These visual insights underscore the need for targeted interventions and policies addressing these critical sociodemographic factors to improve mental health outcomes.

**Fig 1.**
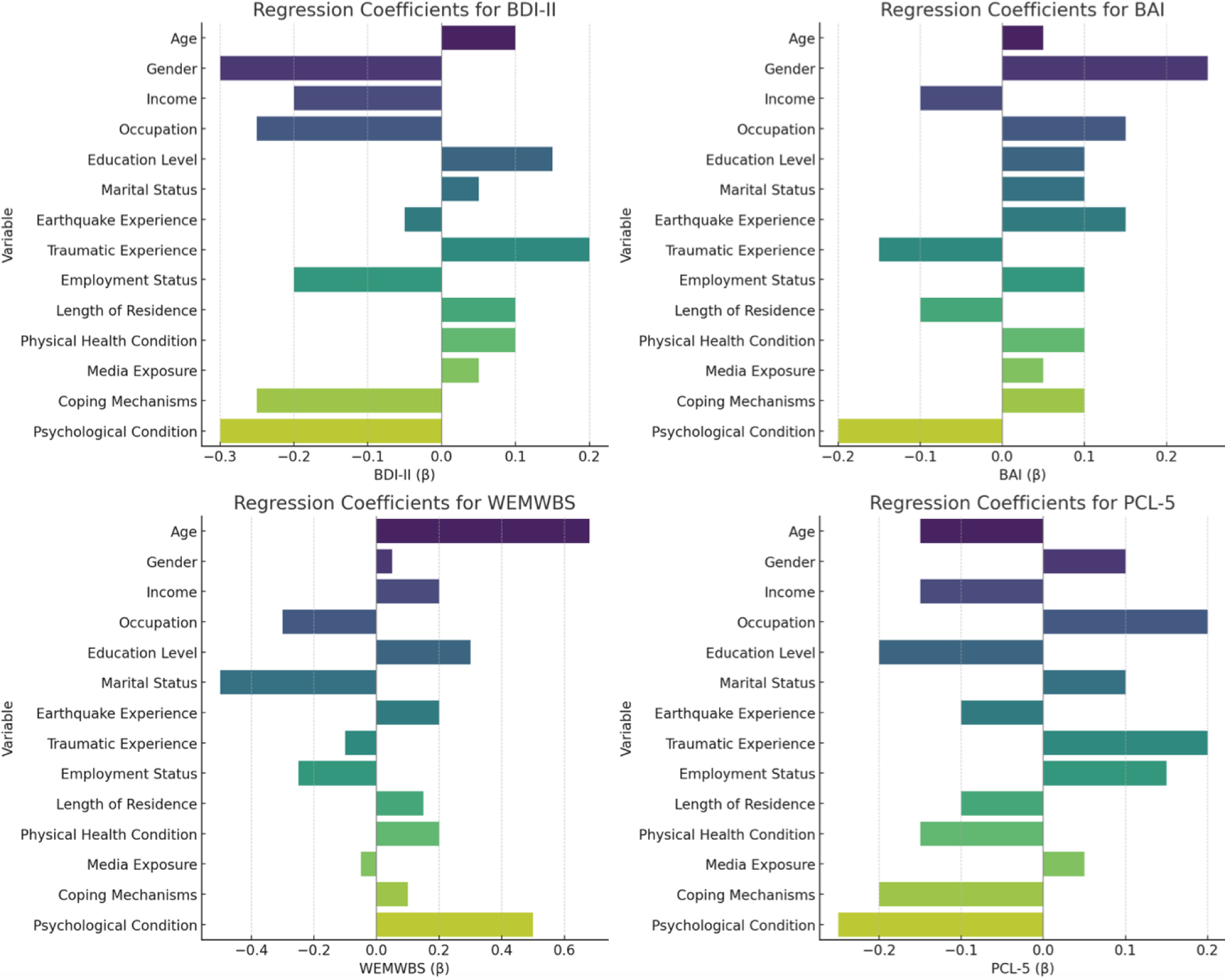
Regression analyses coefficients for psychological scales

## Discussion

The 2023 Kahramanmaraş earthquakes have had a profound impact on the psychological well-being of both direct victims and non-victims. While extensive research has been conducted on the psychological effects of earthquakes on those directly affected [56], this study uniquely focuses on the indirect impact on non-victims, particularly residents of Istanbul. The findings reveal significant psychological distress among non-victims, highlighting the broader societal implications of natural disasters. This discussion aims to contextualize these results within the existing literature, explore the influence of sociodemographic factors and coping mechanisms, and suggest policy implications for comprehensive disaster response and mental health interventions. By examining the psychological aftermath of the earthquakes on non-victims, this study contributes to a more holistic understanding of disaster impacts and underscores the need for inclusive mental health strategies.

Our findings indicate significant psychological impacts on non-victims of the 2023 Kahramanmaraş earthquakes [Table 2], primarily characterized by heightened anxiety and stress levels due to indirect media exposure. This aligns with studies on direct victims of earthquakes, which reveal similarly profound psychological distress [57]. Direct victims faced immediate and tangible stressors such as fear of dying, helplessness, and continuous anxiety about aftershocks, leading to more intense emotional responses and higher PTSD rates [58]. In contrast, our non-victim participants reported distress mainly due to continuous media coverage and perceived threats. The direct exposure experienced by victims resulted in higher helplessness scores and lower problem-focused coping scores compared to those indirectly affected. Social support dynamics also differed significantly between the two groups; non-victims often felt isolated despite high media consumption, possibly due to the lack of direct community engagement, while victims received more immediate practical support [59], though their satisfaction with this support varied. These findings underline the complexity of psychological responses to natural disasters [60], emphasizing the need for tailored mental health interventions [61] that consider both direct and indirect exposures. Victims require immediate and practical support to manage acute stressors [62], whereas non-victims may benefit from interventions aimed at managing media-induced stress and enhancing community support networks [63]. This comparison highlights the broader societal implications of natural disasters and the necessity for comprehensive mental health strategies.

Our findings on the psychological impacts of the 2023 Kahramanmaraş earthquakes on non-victims indicate significant distress primarily driven by indirect media exposure. This mirrors the profound psychological effects documented among direct victims of the 1999 Marmara earthquakes [64]. The Marmara earthquakes led to widespread PTSD and major depression (MD) among survivors, with PTSD prevalence ranging from 8% to 63% and MD from 11% to 42% in various studies [65]. The victims faced immediate stressors such as loss of loved ones, injury, and the destruction of property, which resulted in intense psychological trauma [66]. Similarly, our study found that non-victims of the Kahramanmaraş earthquakes experienced significant anxiety and stress due to continuous media coverage and perceived threats, although their exposure was indirect. In terms of sociodemographic influences, both studies highlight the role of factors like gender [67], income [68], and previous psychiatric history [69] in shaping psychological outcomes. The Marmara earthquake studies noted higher PTSD rates among females, those with lower income [70], and individuals with prior psychiatric issues [71]. Our findings also indicate that non-victims with higher income and education levels reported better mental health, suggesting that these factors provide resilience against disaster-induced psychological distress. The long-term psychological effects observed in both contexts emphasize the need for sustained mental health interventions [72]. The Marmara earthquakes’ victims exhibited persistent PTSD and MD symptoms years after the disaster [73], indicating long-lasting impacts. Similarly, the significant distress reported by non-victims of the Kahramanmaraş earthquakes suggests potential long-term psychological effects if left unaddressed. This parallel underscore the necessity for ongoing mental health support and the development of strategies to mitigate long-term impacts for all affected individuals. Social support dynamics also showed differences between non-victims and direct victims [74]. Non-victims in our study often felt isolated despite high media consumption, possibly due to the lack of direct community engagement. In contrast, direct victims of the Marmara earthquakes received immediate practical support [75], which varied in satisfaction but was crucial for their mental health outcomes. These findings highlight the importance of tailored mental health interventions that address both direct and indirect exposures, ensuring comprehensive disaster response strategies that foster resilience and recovery across affected communities.

Our findings on the psychological impacts of the 2023 Kahramanmaraş earthquakes on non-victims reveal significant distress primarily driven by indirect media exposure. This contrasts with the psychological effects documented among direct victims of other major earthquakes, such as the 1995 Great Hanshin-Awaji earthquake in Japan [76], the 2008 Wenchuan earthquake in China [77], and the 2000 Iceland earthquakes [78]. In the case of the 1995 Great Hanshin-Awaji earthquake, studies like those by Yamamura (2012) have documented long-term declines in subjective well-being among direct victims [79]. The victims faced immediate stressors such as the loss of loved ones and property destruction, leading to high levels of PTSD and major depression [80]. Our study found that non-victims of the Kahramanmaraş earthquakes experienced significant anxiety and stress due to continuous media coverage and perceived threats, despite not being directly impacted by the disaster. This suggests that while the direct victims faced acute trauma, non-victims were still substantially affected through indirect exposure. Similarly, the 2008 Wenchuan earthquake had devastating effects on the psychological well-being of survivors, with studies reporting high levels of PTSD, depression, and anxiety even years after the event [81]. Direct victims of the Wenchuan earthquake faced extreme trauma, including witnessing mass casualties and enduring severe danger [82]. Our findings indicate that non-victims of the Kahramanmaraş earthquakes also reported significant psychological distress, highlighting the far-reaching impacts of such disasters. The long-term psychological effects observed in both contexts underscore the need for sustained mental health interventions. The significant distress reported by non-victims of the Kahramanmaraş earthquakes suggests potential long-term psychological effects if left unaddressed. The Iceland earthquakes in 2000 also provides relevant insights. Bödvarsdóttir and Elklit (2004) found that 24% of direct victims met the criteria for PTSD three months post-disaster [83]. The victims exhibited high levels of anxiety and emotional coping difficulties. In our study, non-victims similarly experienced significant anxiety and stress due to indirect exposure, underscoring the broader societal implications of such events. The Iceland earthquake study noted higher PTSD rates among females and individuals with prior psychiatric issues, which aligns with our findings that sociodemographic factors like gender and income significantly influence psychological outcomes.

In summary, our study highlights the significant psychological impacts of the 2023 Kahramanmaraş earthquakes on non-victims, emphasizing the distress caused by indirect media exposure. This distress mirrors the profound psychological effects documented among direct victims of major earthquakes such as the 1995 Great Hanshin-Awaji earthquake, the 1999 Marmara earthquake the 2008 Wenchuan earthquake, and the 2000 Iceland earthquakes. While direct victims face acute trauma from immediate stressors like loss and destruction, non-victims experience substantial anxiety and stress through continuous media coverage and perceived threats. Both groups require tailored mental health interventions to address their unique needs. The comparisons underscore the complexity of psychological responses to natural disasters and the necessity for comprehensive disaster response strategies that foster resilience and recovery across all affected communities. By addressing the mental health needs of both direct and indirect victims, we can enhance the overall resilience and well-being of societies in the wake of such devastating events.

## Limitations

This study has several limitations that should be considered when interpreting the findings. First, the cross-sectional design of the study only captures a snapshot of the psychological impact at a single point in time, limiting our ability to assess changes in mental health over the long term. Longitudinal studies are needed to understand the persistence and evolution of psychological distress among non-victims.

Second, the reliance on self-reported measures may introduce bias, as participants might underreport or overreport their symptoms due to social desirability or recall bias. Additionally, while the sample size was substantial, it may not be fully representative of the broader population of non-victims across Turkey, as the majority of participants were from Istanbul.

Third, the study did not account for potential confounding variables such as pre-existing mental health conditions, which could influence the psychological outcomes observed. Although we excluded participants with known psychiatric histories, undiagnosed conditions may still have affected the results.

Fourth, the study focused primarily on media exposure as the main factor influencing psychological distress. Other factors, such as personal connections to victims or secondary stressors related to the earthquakes (e.g., economic impact, disruptions in daily life), were not extensively examined and could contribute to the observed effects.

## Conclusion

The 2023 Kahramanmaraş earthquakes have highlighted the far-reaching psychological impacts of natural disasters, extending beyond the immediate zones of physical destruction to affect individuals indirectly exposed through media and social networks. Our study reveals that non-victim, particularly those in Istanbul, experienced significant levels of anxiety, stress, and symptoms of PTSD due to continuous exposure to distressing media coverage. These findings underscore the broader societal implications of such disasters and the necessity for mental health interventions that address the needs of both direct victims and non-victims.

Comparisons with studies on direct victims of the 1995 Great Hanshin-Awaji earthquake, the Marmara earthquake 1999, the 2008 Wenchuan earthquake, and the 2000 Iceland earthquakes further illustrate the profound psychological distress caused by immediate exposure to traumatic events. While direct victims endure acute stressors such as loss of life and property, non-victims suffer from a pervasive sense of vulnerability and fear, driven by media consumption and perceived threats. Both groups require tailored mental health interventions to mitigate long-term psychological effects and foster resilience.

Our research emphasizes the importance of considering sociodemographic factors, such as age, gender, income, and education, which influence psychological outcomes. Higher income and education levels were associated with better mental health among non-victims, suggesting that these factors provide resilience against disaster-induced distress. The study also highlights the need for continuous mental health support and comprehensive disaster response strategies that include psychological first aid, ongoing counseling services, and community support programs.

In conclusion, the psychological impact of the 2023 Kahramanmaraş earthquakes on non-victims highlights the necessity for inclusive mental health strategies that address both direct and indirect exposures to natural disasters. By understanding and addressing the broader societal effects, we can enhance the resilience and well-being of communities, ensuring a more robust recovery process in the aftermath of such catastrophic events.

## Supplemental Material Statement

All supplemental materials related to this study are available on Figshare at the following link: https://figshare.com/s/23cf21b898e522a5bb07

## Data Availability

https://figshare.com/s/23cf21b898e522a5bb07

## Acknowledgements

We would like to thank Sevil Akman, Vuslat Cihanlıgil, Ülkü Karaşlıoğlu, and Emre Orhanlı for their efforts in collecting the data. Additionally, we extend our gratitude to all the participants for their valuable contributions to this study.

## Author Contribution

### Conceptualization

Metin Çınaroğlu, Eda Yılmazer **Methodology**: Gökben Hızlı Sayar, Zeynep Alpugan **Collecting the data:** Metin Çınaroğlu, Eda Yılmazer **Writing original draft:** Metin Çınaroğlu, Eda Yılmazer **Supervision:** Gökben Hızlı Sayar

### Data Analyses and visulizations

Zeynep Alpugan

